# Village-level surveillance of neonatal disease with integrated real-time dashboards and quality-control in Uganda

**DOI:** 10.64898/2026.07.21.26358401

**Authors:** Joseph N. Paulson, Andrew J. Whalen, Starlin Tindimwebwa, Justice Hansen, Davis Natukwatsa, Kalifuba Steven, Moses Ochora, Ronnie Mulondo, Edith Mbabazi Kabachelor, Kamron Ramelmeier, Brian Kaaya Nsubuga, Philip O. Omadi, Joshua Magombe, Carmit Cohen, Norbert Muzahura, Justin Onen, Peter Ssenyonga, James R. Broach, Sarah U. Morton, Marwan Osman, Moses Joloba, Edgar Kigozi, Andrew Katabalwa, Julian Apako, Hilda Amutuhaire, John Baptist Tumuhairwe, Agatha Kayemba, Josephine Namyalo, Henry Masengere, Harriet Nambuya, Agatha Namutosi, Sophia Kasuswa, Emma Omo, Ivan Tibenkana, Alfred Yayi, Joseph Muvawala, William Nadiope, Abraham Muwanguzi, Elias Kumbakumba, Jessica E. Ericson, Steven J. Schiff

## Abstract

**Introduction:** Neonatal mortality remains disproportionately high in sub-Saharan Africa, where an estimated 27 neonatal deaths per 1,000 live births occur annually. Infections, including sepsis and meningitis, account for a substantial proportion of these deaths, while neural tube defects (NTDs) contribute significantly to both neonatal mortality and long-term disability. Existing surveillance systems in the region are predominantly facility-based, missing the substantial proportion of births and deaths that occur in the community. Population-based surveillance platforms that capture community-level data are urgently needed to generate accurate incidence estimates, identify modifiable risk factors, and guide evidence-based interventions.

**Cohort Description:** The Consortium to Reduce Infant Mortality (CONRIM) is a multi-institutional partnership among Ugandan physicians and scientists, Yale University, Penn State University, Boston Children’s Hospital/Harvard Medical School, and Uganda’s National Planning Authority. CONRIM conducts prospective, community-based neonatal surveillance within the Busoga Kingdom in eastern Uganda. A network of 813 trained Village Health Team members conducts household-level visits using a structured Open Data Kit (ODK)-based mobile questionnaire to capture every birth, assess for danger signs of possible serious bacterial infection (pSBI), screen for NTDs, and record maternal nutrition and folic acid use, water, sanitation and hygiene (WASH) conditions, and health care utilization.

**Findings to Date:** Since surveillance began in June 2025, the platform has registered approximately 22,200 household submissions and over 5,700 newborn encounters across the Jinja District (population 660,000). Early data have identified higher than expected rates of infants with NTDs including encephalocele and spina bifida; documented folic acid non-use in before and during most pregnancies; characterized WASH conditions in birthplaces; and mapped geospatial hotspots of neonatal infection risk in northeastern rural subcounties. Prospective 28-day follow-up of all live births has demonstrated a neonatal mortality rate of 21.5 per 1000 live births. A real-time data quality monitoring system with 21 automated quality control flags maintains a 99% clean-record rate.

**Future Plans:** Ongoing and planned activities include laboratory-based confirmation of neonatal sepsis via blood culture and cerebrospinal fluid analysis with polymerase chain reaction capacity, portable neuroimaging for NTDs, environmental sampling, genomic studies of folate metabolism pathway genes, linkage with facility-based records at Jinja Regional Referral Hospital and Mulago National Referral Hospital, and community-level interventions informed by surveillance findings.

**Key Messages:** *What is already known on this topic:* Neonatal mortality remains disproportionately high in sub-Saharan Africa, with sepsis and neural tube defects (NTDs) among the leading preventable causes. Existing surveillance systems are predominantly facility-based and fail to capture births, deaths, and environmental exposures occurring at the community level. Emerging approaches in digital health, geospatial analytics, and pathogen genomics have demonstrated potential to enhance infectious disease surveillance, but these have rarely been integrated into population-based neonatal monitoring systems in low-resource settings.

*What this study adds:* The **Consortium to Reduce Infant Mortality (CONRIM)** is a multidisciplinary initiative designed to develop scalable, population-based systems for understanding and reducing neonatal mortality through integrated epidemiologic, environmental, and biologic data. This paper describes one implementation of the CONRIM framework in the Busoga Kingdom of eastern Uganda, where a network of 813 trained Village Health Team members conducts longitudinal, community-based surveillance of births, neonatal outcomes, NTDs, maternal nutrition (including folic acid use), water, sanitation and hygiene (WASH) conditions, and care-seeking behaviour. This implementation integrates:

- real-time digital data capture with automated quality control,
- geospatial information systems (GIS) and remote sensing to characterize environmental risk factors,
- population-level genomic and metagenomic sampling to investigate host and pathogen factors, and
- a One Health framework linking human, animal, and environmental exposures. Early findings highlight high data completeness, geospatial clustering of neonatal infection risk, low preconception folic acid use, and identification of NTD cases not captured by facility-based systems.

*How this study might affect research, practice, or policy:* This study demonstrates the feasibility of implementing a community-based, real-time neonatal surveillance system within an existing community health worker network in a low-resource setting. By integrating geospatial, genomic, and environmental data within a unified platform, the CONRIM framework enables more precise identification of drivers of neonatal morbidity and mortality. The approach supports targeted public health interventions, including geographically informed infection control strategies, improved referral pathways, and evidence generation for folic acid fortification policies. More broadly, CONRIM provides a scalable infrastructure for future interventional studies and precision public health strategies aimed at reducing neonatal mortality.

## Introduction

### The Burden of Neonatal Mortality in Sub-Saharan Africa

Globally, an estimated 2.3 million neonates die within their first 28 days of life each year, accounting for nearly half of all deaths in children under five years old (Gu et al., 2026, https://www.who.int/news-room/fact-sheets/detail/newborn-mortality). Sub-Saharan Africa bears a disproportionate share of this burden, with a regional neonatal mortality rate of approximately 27 per 1,000 live births, more than double the global average (https://www.who.int/news-room/fact-sheets/detail/newborn-mortality). Uganda, despite substantial progress in reducing child mortality over recent decades, continues to report a neonatal mortality rate of approximately 19 per 1,000 live births, with progress in neonatal survival lagging behind improvements in post-neonatal child mortality (Mejía-Guevara et al., 2019).

The three major causes of neonatal death globally are birth complications including prematurity and birth asphyxia, congenital malformations, and infections(Liu et al., 2016). Neonatal sepsis and meningitis are estimated to cause 15-25% of all neonatal deaths worldwide, with the burden concentrated in low- and middle-income countries (LMICs) where access to clean delivery environments, postnatal care, adequate diagnostic studies and timely treatment remains limited (Jena & Jaldo, 2025). Hydrocephalus is a frequent sequela of meningitis, often requiring surgical intervention (Warf, 2005). In sub-Saharan Africa, the incidence of community-acquired neonatal infections is impacted by non-sterile cord care practices, delayed medical evaluation, antimicrobial resistance, and limited access to microbiological diagnostic services (Jena & Jaldo, 2025)(Hopp, 2017).

Neural tube defects are among the most common and severe congenital malformations and represent a further under-recognised contributor to neonatal mortality and disability in the region (O’Leary et al., 2026). NTDs include anencephaly, which is invariably fatal, spina bifida and encephalocele, which have significant morbidity and mortality even with prompt surgical intervention. Many NTD-affected pregnancies result in stillbirth or early neonatal death in the community and are not included in hospital-based incidence measures, leading to substantial underestimation of the true burden of these conditions. The relationship between NTD prevalence and inadequate folic acid supplementation before and during early pregnancy is well established, yet population-level data on both folic acid uptake and NTD prevalence remain sparse across most of sub-Saharan Africa (Katunzi et al., 2025).

### Gaps in Surveillance

The vast majority of existing neonatal health data in sub-Saharan Africa originate from facility-based sources: hospital registries, demographic and health surveys, and health management information systems (Nkurunziza et al., 2026). While these data sources are valuable, they systematically miss births and deaths occurring in the community. In many parts of Uganda, a substantial proportion of deliveries take place at home or are attended by traditional birth attendants (TBAs), and neonates who fall ill or die may never present to a health facility (Montagu et al., 2011). Facility-based data therefore underestimate the true incidence of neonatal morbidity and mortality and are subject to referral and ascertainment bias.

Population-denominator data suitable for calculating community-level incidence rates are particularly scarce. Without knowing the true birth rate, it is not possible to estimate incidence and compare risk across areas, identify high-burden populations, or evaluate the impact of interventions. Furthermore, there is limited integration of clinical data with the environmental, nutritional, and demographic determinants that shape neonatal outcomes. Addressing these gaps requires surveillance platforms that are community-based, population-defined, and capable of capturing multiple data streams simultaneously.

### The CONRIM Initiative

The Consortium to Reduce Infant Mortality (CONRIM) was established as a multidisciplinary, multinational effort to identify key risk factors for neonatal mortality coupled with targeted prevention efforts to ultimately reduce infant mortality globally. The prototype surveillance system to address these challenges was developed and deployed as a comprehensive, community-based neonatal surveillance platform in eastern Uganda. CONRIM is a multi-institutional collaboration among Ugandan physicians and scientists, Yale University, Penn State University, Boston Children’s Hospital / Harvard Medical School, and Uganda’s National Planning Authority (NPA), with close partnership with the Busoga Kingdom traditional governance structures. The consortium’s mission is to integrate clinical, environmental, and demographic data streams to generate population-level evidence on the neonatal disease burden, identify modifiable risk factors, and guide evidence-based policy and interventions to reduce preventable neonatal mortality.

The Busoga Kingdom in eastern Uganda was selected as the study setting for several reasons: its population of over four million people provides a large denominator for incidence estimation; it encompasses a mix of urban, peri-urban, and rural communities with varying access to health services; it has an established Village Health Team (VHT) infrastructure that provides a foundation for community-based data collection; and the Busoga Kingdom’s traditional governance structures, led by the Kyabazinga (King), offer a mechanism for community engagement and stakeholder buy-in that is invaluable for sustainable surveillance and the implementation of public health programs aimed at reducing infant mortality.

### Aims of This Paper

This paper presents the structure and application of the CONRIM neonatal surveillance platform. Specifically, we aim to: (1) describe the design, implementation, and data infrastructure of the CONRIM community-based surveillance system; (2) present the baseline characteristics of the cohort enrolled to date; (3) report preliminary descriptive findings on neonatal mortality, NTDs, and key maternal-infant risk factors; and (4) outline the platform’s capacity for ongoing and future research, including laboratory-based sepsis confirmation, neuroimaging, and genomic studies as well as ongoing surveillance for the effectiveness of public health interventions.

### Cohort Description

#### Study Setting

The CONRIM surveillance platform operates within Jinja District of the Busoga Kingdom in eastern Uganda. The Busoga Kingdom has a total population estimated at over four million, with an estimated annual birth rate of 35-40 per 1000 population. Jinja District alone has an estimated annual birth cohort exceeding 7,500, of which approximately 6,000 facility-based deliveries are recorded at Jinja Regional Referral Hospital (JRRH).

The health system in the Busoga Kingdom follows Uganda’s tiered structure, with Health Centre II facilities at the parish level, Health Centre III at the subcounty level, Health Centre IV (capable of performing caesarean sections) at the county level, and JRRH serving as the regional referral facility (Muluya et al., 2025). Traditional birth attendants remain active in some communities, particularly in rural areas, and home deliveries without any attendant also occur (Kagoya et al., 2026). VHTs, comprising community health volunteers appointed by their local councils, form the backbone of the community health outreach system and serve as the primary data collection workforce for CONRIM (Lee et al., 2025).

#### Study Design

CONRIM is a prospective, population-based, community-level cohort surveillance study. The study employs continuous enrolment: all households within the surveillance area that have a current pregnancy or a recent delivery are eligible for inclusion. There are no exclusion criteria at the household level; the target is universal coverage of all births within the catchment area. The surveillance platform began data collection in June 2025, with enrollment ongoing.

#### Ethics and Governance

The study was approved by the Yale University Institutional Review Board (protocol #2000040126, approved 30 July 2025) and has received local ethical review and administrative clearance from the Research Ethics Committee of the Mbarara University of Science and Technology, the Uganda National Council for Science and Technology, and relevant district authorities (protocol #MUST-2024-1657, approved 8 August 2024). Informed consent is obtained from all caregivers participating in the study survey and prior to collection of any personally sensitive information regarding home life and neonatal care practices. Consent may be obtained via electronic signature, or written signature (captured electronically via photograph of the signed consent form), following discussion of the study in the participant’s local language by trained VHTs. Study participation cards bearing a unique study identification number are issued to all enrolled households. Weekly consent rates have approached 97-100% in recent data collection periods.

### Participant Recruitment and Data Collection

#### Village Health Teams

A network of 813 VHTs serves as the primary data collection workforce across a single district. VHTs are community health volunteers who reside in the villages they serve and are appointed by local government structures. For CONRIM, VHTs undergo structured training in the use of the ODK Collect mobile application, recognition of neonatal danger signs based on World Health Organization (WHO) and Uganda Ministry of Health criteria, NTD visual screening, the informed consent process including training in the responsible conduct of research, and data quality standards. Training is reinforced through monthly field supervision visits, refresher sessions, WhatsApp-based support groups with visual aids, and community meetings with VHT leadership at the subcounty and parish levels. Participating VHTs are paid a nominal stipend on a monthly basis.

VHT performance is monitored through an automated quality control scorecard system that tracks submission rates and error frequencies, and flags patterns at the individual VHT level. VHTs with high error rates or low submission volumes are identified for targeted retraining during field supervision visits.

#### Household Visits and Enrolment Flow

VHTs conduct systematic household visits within their assigned villages to identify pregnant women and recent deliveries with regular visits to every village. Upon identifying an eligible household, the VHT explains the study, obtains informed consent, and administers the structured questionnaire using the ODK Collect application on an Android mobile device. If the infant is alive, the full questionnaire is administered, covering infant health status, danger signs, birth defects, delivery details, maternal health, and household conditions. If the infant has died, an abbreviated form is administered that captures available information on the circumstances and possible cause of death, incorporating verbal autopsy elements. The study flow is depicted in Figure 1.

**Figure 1.**
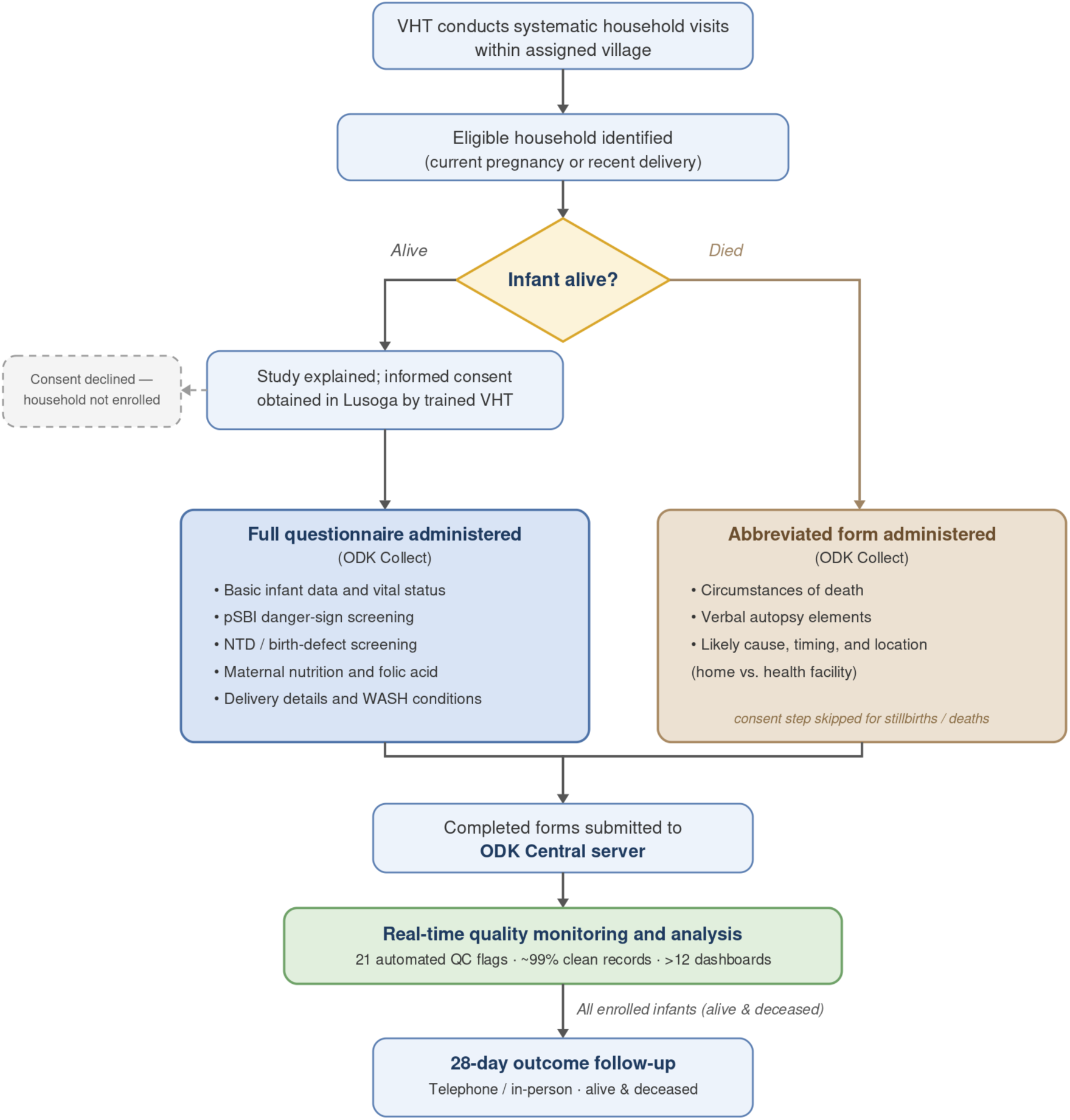
CONRIM study flow chart. VHTs identify eligible households and administer the structured questionnaire via the ODK Collect mobile application. If the infant is alive, the full questionnaire is administered; if the infant has died, an abbreviated form with verbal autopsy elements is used. Completed forms are submitted to the ODK Central server for real-time quality monitoring and analysis.

**Figure 2.**
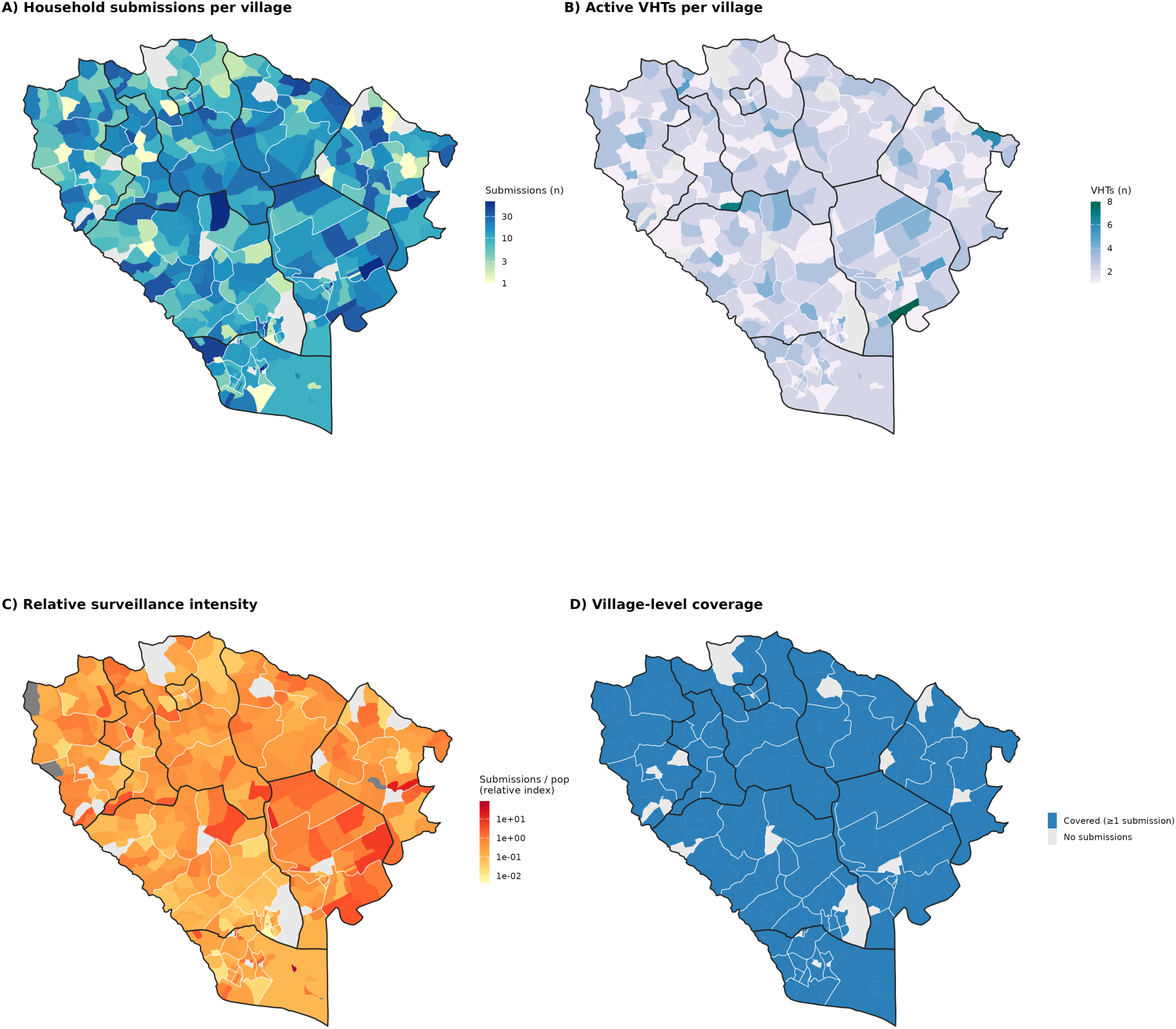
Village-level VHT surveillance coverage across the CONRIM catchment, Jinja District and Jinja City, eastern Uganda. Each polygon represents one of 417 villages; bold black lines delineate the 9 subcounties and thin white lines the 60 parishes. Submission records were joined to village polygons using the surveyor-selected village identifier. (A) Total household submissions per village (color scale log-transformed; range 0-373). (B) Number of distinct active Village Health Team (VHT) members submitting from each village. (C) Relative surveillance intensity, defined as submissions per unit population; values are shown on a log scale as a relative index rather than a literal per-capita rate, because the available population layer is a scaled WorldPop-derived estimate. (D) Binary village-level coverage, distinguishing villages with at least one recorded submission from those with none. Across the catchment, 389 of 417 villages (93%) recorded ≥1 submission during the surveillance period; villages with no submissions are shown in grey in all panels.

#### Data Collection Instruments

Data are captured using the ODK Collect mobile application, with completed forms transmitted to an ODK Central server. The structured questionnaire comprises eight integrated case reported forms (CRFs):

### Basic Infant Data, Infection Screening, and Birth Defect Screening CRFs

These core forms capture infant demographics (sex, date of birth, birth weight, gestational age), vital status, danger signs consistent with possible serious bacterial infection (pSBI) (fever, convulsions, poor feeding, fast breathing, chest indrawing, lethargy, umbilical redness or discharge, skin pustules), and screening for NTDs and other birth defects (back swelling, encephalocele, microcephaly, hydrocephalus).

In addition to infant health data, the questionnaire captures maternal and household-level variables including folic acid supplementation (timing relative to pregnancy, dose, and source), antenatal care attendance, mode and place of delivery, breastfeeding practices, water source (differentiated by rainy and dry season), latrine access, handwashing practices, and substances applied to the umbilical cord.

Finally, a centralized study team calls each enrolled family after the neonate reaches 28 days of age to complete a follow-up outcomes assessment using a semi-structured interview.

### Follow-Up Outcomes

Captures 28-day follow-up data via telephone and in-person visits, including vital status, health status including resolution of any pSBI symptoms, healthcare utilisation, and referral completion.

### Diagnostic Definitions

#### Possible Serious Bacterial Infection (pSBI)

pSBI is defined based on WHO and Uganda Ministry of Health criteria - an infant with a “neonatal danger sign” is considered to have a pSBI. These criteria are designed to identify neonates who require urgent evaluation by a health professional and include: fever or hypothermia, convulsions, poor feeding or inability to suck, fast breathing, severe chest indrawing, lethargy or reduced movement, umbilical redness or purulent discharge, and skin pustules. VHTs are trained to assess and record the presence or absence of each danger sign during the household visit. pSBI are classified as early-onset (<72 hours from birth) or late-onset (≥72 hours from birth) where the timing of symptom onset can be ascertained. The neonatal period is defined as the first 28 days of life.

#### Neural Tube Defects

NTD screening is conducted by VHTs through visual inspection of the infant and caregiver report. NTDs captured include protuberant back tissue (consistent with spina bifida or myelomeningocele), encephalocele, microcephaly, and anencephaly. Anencephaly is often captured within stillbirth reports, as affected pregnancies frequently result in fetal or early neonatal death. Infants with suspected NTDs are referred for confirmation through follow-up by the CONRIM field team and, for surgical intervention, referral to CURE Children’s Hospital of Uganda in Mbale or Mulago National Referral Hospital (MNRH) in Kampala. Confirmed diagnoses of NTD are tracked through the follow-up system, and surgical outcomes are recorded where applicable.

#### Follow-Up

All live births enrolled in the surveillance platform have follow-up at 28 days of age. For infants identified as sick at initial assessment, intensified follow-up is conducted at approximately 28 days. Follow-up is conducted by trained study staff via telephone, using a scripted interaction delivered in the participant’s language. The follow-up script addresses infant vital status, relevant pBSI symptom resolution or progression, healthcare utilisation since initial enrolment, referral completion for those previously referred, and any new concerns. When the infant has died, verbal autopsy elements are incorporated into the follow-up to ascertain the likely cause, timing and location (home vs. health facility) of death. When the caregiver for the infant cannot be reached, the infant is classified as unreachable and documented accordingly.

### Clinical and Environmental Diagnostic Capacity

A central objective of the CONRIM platform is to move beyond syndromic surveillance of pSBI based on clinical symptoms toward laboratory-confirmed diagnosis of neonatal infections and robust etiological identification of NTDs. To support this initiative, an integrated framework of laboratory and diagnostic workstreams is being developed to provide accurate pathogen detection and antimicrobial susceptibility testing.

### Neonatal Sepsis and Post-Infectious Hydrocephalus Laboratory Confirmation

Neonates presenting to JRRH with signs of sepsis or meningitis, or to Mulago Hospital (MNRH) with hydrocephalus (either congenital or post-meningitic), who enroll in the study, will complete additional data collection forms relevant to their clinical condition. Blood culture and cerebrospinal fluid (CSF) cultures for neonates with suspected sepsis or meningitis are performed at both sites being implemented at JRRH and Mulago HospitalMNRH. At MNRH Mulago Hospital, patient samples collected at MNRH from this site are processed analyzed at the Makerere University Biomedical Research Centre (MakBRC), a College of American Pathologists (CAP)-accredited, ISO 15189-certified clinical laboratory located on the same campus in Kampala, Uganda. All samples undergo an extended culture protocol, incorporating both liquid media (blood culture bottles) and solid media, including Chocolate Agar with polyvitex, Columbia CNA Agar, Brain Heart Infusion Agar, MacConkey Agar, and Sabouraud Dextrose Agar. Gram stain is performed on all CSF specimens and on all positive blood culture specimens bottles. Growing bacterial colonies are identified using Matrix-Assisted Laser Desorption/Ionization Time-of-Flight Mass Spectrometry (MALDI-TOF MS) (Bruker Daltonics) or Vitek 2 (bioMérieux) at MakBRC and JRRH, respectively. Antimicrobial susceptibility is determined using the Kirby-Bauer disk diffusion method as described in the guidelines of the Clinical and Laboratory Standards Institute (CLSI). Real-time Polymerase Chain Reaction (PCR) capacity is available at both sites being established for pathogen detection, with an initial focus on *Paenibacillus thiaminolyticus*, a fastidious pathogen identified in Uganda as a major cause of neonatal sepsis and post-infectious hydrocephalus in infants (Paulson et al 2020; Morton et al 2023; Ericson et al 2023). The existing microbiology laboratory has been strengthened to ensure high-quality diagnostics, emphasizing good laboratory practices and robust quality control measures through staff training and external audits. In addition, a dedicated molecular biology laboratory has been set up within the clinical laboratory at JRRH.

Forms that would cover these topics and some of which are part of future work include:

### Hydrocephalus Clinical CRF

Detailed clinical assessment for infants identified with suspected hydrocephalus.

### Laboratory CRF

Records results from blood cultures, cerebrospinal fluid analysis, Gram stain, antimicrobial susceptibility testing, and PCR-based pathogen identification.

### Imaging CRF

Captures neuroimaging findings from portable magnetic resonance imaging (pMRI) assessment of NTDs and hydrocephalus.

### Surgical CRF

Documents surgical interventions, including procedures performed at CURE Children’s Hospital of Uganda or MNRHMulago National Referral Hospital for NTD repair.

### Environmental Sampling

Environmental sampling has been initiated to characterise household- and community-level exposures that may contribute to neonatal infection risk. Samples included interior wall swabs, soil, and diverse water sources such as boreholes, rivers, household taps, wells, and stored water in jerrycans. Fecal specimens were collected from backyard livestock, including pigs, chickens, cows, goats and ducks. In addition, raw food items—including locally sourced fish and unpasteurized cow’s milk—were sampled to evaluate potential microbial exposure pathways.

Real-time PCR assays are being performed on these samples to detect microbial pathogens, with an initial focus on *P. thiaminolyticus*. These data are being integrated with satellite-derived environmental variables including land cover classification, watershed boundaries, and a multidimensional poverty index to build a comprehensive environmental risk factor profile for neonatal morbidity and mortality.

### Genomic Studies

Planned genomic studies will examine variants in folate metabolism pathway genes that may contribute to NTD susceptibility in this population, as well as ancestral admixture genomic variations at the population that contribute to neonatal infection risk and outcomes.

## Data Management and Quality Assurance

### Data Infrastructure

Data flow from the point of collection to analysis through a structured pipeline. VHTs submit completed questionnaires via the ODK Collect mobile application to the ODK Central server. Data are then processed and displayed through a suite of more than 12 real-time interactive dashboards covering household enrolment, newborn health, infant demographics, maternal nutrition, geographic analysis, WASH and environmental conditions, consent monitoring, follow-up management, and data quality metrics (Figure 3). These dashboards are accessible to the study team and are used for weekly data review meetings. Additionally, an automated system generates weekly PowerPoint summary reports for stakeholder review. A separate REDCap database has been established for laboratory specimen tracking at the hospital sites (JRRH and MNRH).

**Figure 3.**
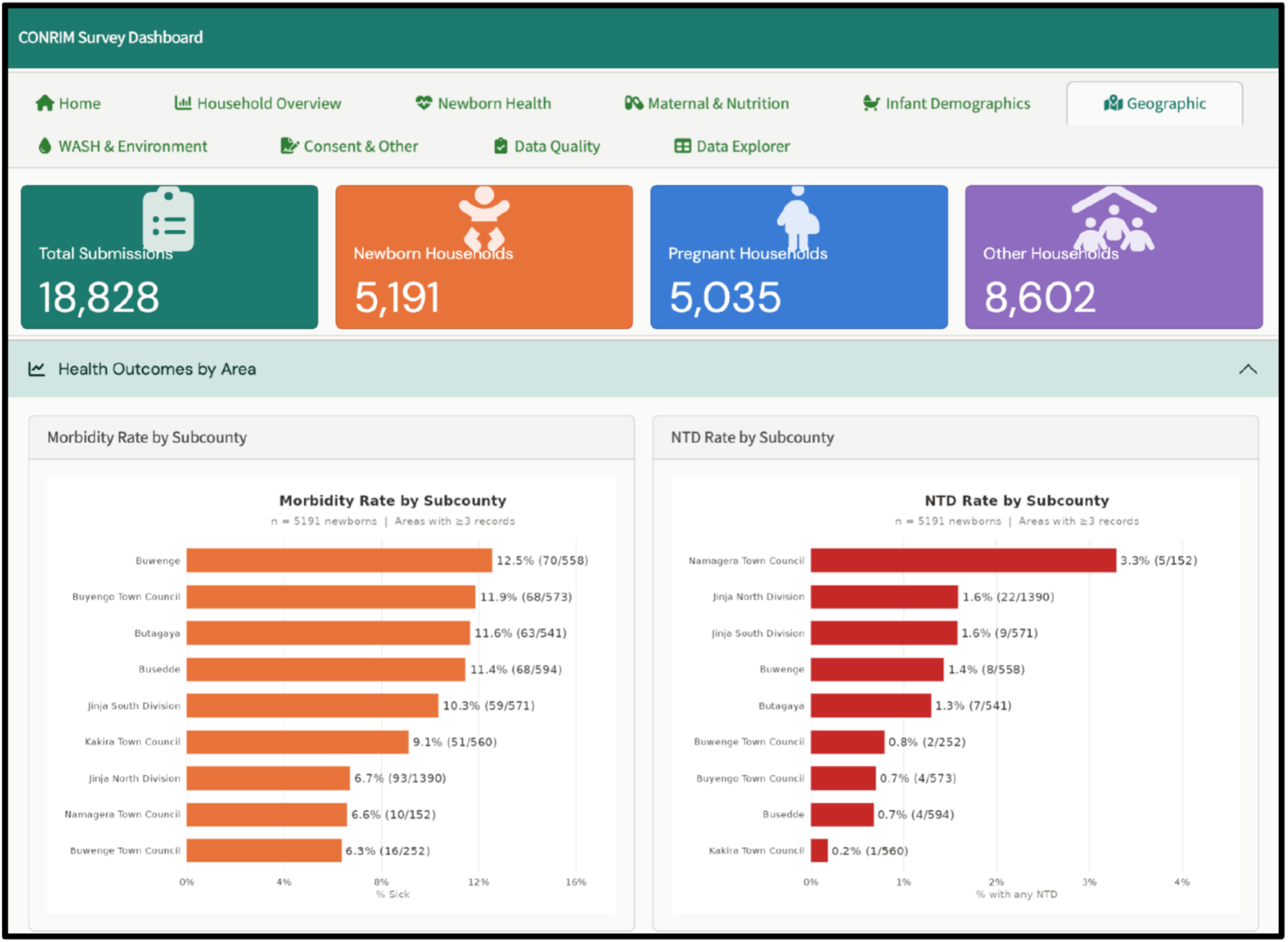
CONRIM real-time data dashboards. Representative screenshots showing data quality monitoring morbidity and NTD rates by subcounty.

### Quality Control System

Data quality is monitored through a multi-layered system combining automated and manual review. Twenty-one automated quality control flags are applied to each submission upon receipt, covering categories including possible duplicate entries (matching date of birth, village, sex, and VHT), date inconsistencies, implausible values (such as birth weight exceeding 5 kg with normal vaginal delivery), and missing required fields. The CONRIM Data Quality Monitor dashboard provides real-time visibility into the number of records checked, records with issues, total quality control flags triggered, and the average number of flags per record.

Over the surveillance period, the weekly error rate has declined from approximately 12% in the initial months to under 2% in recent months, with the overall proportion of clean records currently at 99%. This improvement reflects the iterative refinement of VHT training, questionnaire design, and the implementation of VHT-level performance scorecards and flag heatmaps that identify individual VHTs with high error rates for targeted retraining. A manual review workflow complements the automated system: flagged records are entered into a query tracker, reviewed by field supervisors, and resolved through direct communication with the responsible VHT, with a target response time of 48 hours. Weekly data quality meetings with the field team review outstanding queries, error trends, and process improvements.

### Data Governance

Data access and governance are being defined in a CONRIM PI Charter, which establishes distinct access levels for operational data, analytic datasets, and publication-ready datasets. Cross-site analyses are proposed through a structured process and reviewed for scientific merit, overlap with ongoing work, feasibility, compliance, and authorship implications. Sharing of consortium data or materials outside the participating group requires appropriate scientific and institutional approval.

### Findings to Date

#### Enrolment and Coverage

As of July 2026, the CONRIM surveillance platform has received approximately 22,196 total household submissions. Of these, 5,523 are newborn households (households with a recently delivered infant). Weekly submission volumes have shown a sustained upward trend over the surveillance period, reflecting both growing VHT participation and improved community acceptance of the study.

#### Infant Demographics

Among the 5,523 newborns enrolled during the reporting period, 2,873 were male and 2,650 were female. The distribution of birth weights showed 621 infants with low birth weight (<2.5 kg), with 9.3% of the population having a birthweight z-score below −2 (and 1.6% below −3), relative to a fixed term reference birthweight of 3.3 kg (SD 0.5 kg). Gestational age assessment, based on maternal recall of the last menstrual period, classified the majority of births as term, with a smaller proportion reported as preterm. A notable number of initial “don’t know” responses for gestational age were resolved through follow-up to confirm term births (Table 1).

**Table 1.** Baseline characteristics of enrolled newborns (N = 5,523). Values are n (%). Sub-category percentages are of the total; birth term and delivery categories may not sum to 100% owing to missing or “don’t know” responses.

| Characteristic | N (%) |
| --- | --- |
| **Total newborns** | **5,523** |
| **Sex** |  |
| Male | 2,873 (52.0%) |
| Female | 2,650 (48.0%) |
| Low birth weight (<2.5 kg) | 623 (11.3%) |
| **Birth term** |  |
| Term | 4,997 (90.5%) |
| Preterm | 440 (8.0%) |
| Don't know | 86 (1.6%) |
| **Type of delivery** |  |
| Vaginal delivery | 4,693 (85.0%) |
| Caesarean section | 830 (15.0%) |
| **Location of birth** |  |
| Health centre | 3,125 (56.6%) |
| Hospital | 1,767 (32.0%) |
| Home | 250 (4.5%) |
| Private facility | 193 (3.5%) |
| Traditional birth attendant | 147 (2.7%) |
| Other | 41 (0.7%) |

#### Delivery and Birthplace

Among 5,523 newborns for whom delivery data were available, 85.0% were delivered by vaginal delivery and 15.0% by caesarean section. The observed caesarean section rate exceeds the Uganda national average of approximately 6%, likely reflecting the concentration of higher-risk pregnancies at facility-based deliveries captured in the surveillance, as well as regional variation in surgical capacity. The majority of births occurred at health centres (3,125; 56.6%) or hospitals (1,767; 32.0%), with smaller proportions at home (250; 4.5%), a private facility (193; 3.5%), with a traditional birth attendant (147; 2.7%), or at another location (41; 0.7%). The relatively low proportion of home births may reflect both successful national policies promoting facility-based delivery in Jinja District and potential underreporting of community births in the early phase of surveillance.

#### Neonatal Morbidity and Mortality

At the time of initial VHT assessment, 98.4% of enrolled newborns were alive and 1.6% had died. Among living infants, 9.5% were reported as having one or more danger signs consistent with pSBI. The most frequently reported danger signs included fever and fast breathing. Infants with pSBI were stratified by timing of onset where ascertainable, distinguishing early-onset infections (<72 hours) potentially reflecting maternal or intrapartum transmission from late-onset infections (≥72 hours) more likely reflecting community-acquired or nosocomial sources.

Of 5,708 babies who had reached 28 days of age during the reporting period, 66.6% had been followed up. Follow-up completion rates are expected to improve as the study’s telephone follow-up system reaches full operational capacity. Among deaths reported, causes included prematurity, birth asphyxia, and suspected sepsis, though formal verbal autopsy classification is still being standardised (Table 2).

**Table 2.** Neonatal (28-day) mortality among enrolled newborns. Denominators reflect infants who had reached 28 days by the reference date (18 July 2026) and follow-up completeness; ranges and 95% CIs are Clopper-Pearson. NMR, neonatal mortality rate.

| Measure | Value |
| --- | --- |
| Newborns in mortality cohort | 5,741 |
| Reached day 28 (by reference date) | 5,708 |
| Followed up (of those reaching 28 d) | 3,801 |
| Died ≤28 d (numerator) | 80 |
| Survived to 28 d (incl. died later) | 3,648 |
| Known 28-day outcome (denominator) | 3,728 |
| Loss to follow-up | 1,386 |
| Dead, date unknown | 0 |
| Vital status unknown | 571 |
| Uncertain (bounds only) | 1,957 |
| Not yet 28 d old (pending) | 26 |
| Stillbirths (excluded) | 29 |
| Excluded — invalid dates | 1 |
| 28-day survival, complete-case | 97.9% [64.2%-98.6%] |
| Complete-case NMR (per 1,000 known) | 21.5 |
| 95% CI (Clopper-Pearson) | 17.1-26.6 |
| NMR per 1,000 live births (bounded) | 14.1-358.3 |
| NMR incl. stillbirths (per 1,000) | 27.5 |
| 95% CI (Clopper-Pearson) | 22.5-33.2 |
| NMR incl. stillbirths, bounded | 18.0-360.9 |
| Stillbirth rate (per 1,000 births) | 5.1 |

Across the surveillance cohort, 5,741 newborns were recorded, of whom 29 were stillbirths (a stillbirth rate of 5.1 per 1,000 births). Among the 3,728 infants with a known 28-day outcome, 80 had died by day 28, corresponding to a neonatal mortality rate (NMR) of 21.5 per 1,000 live births (95% CI 17.1-26.6); accounting for follow-up completeness, the rate is bounded between 14.1 and 358.3 per 1,000. When stillbirths are additionally counted as events, the combined rate is 27.5 per 1,000 (95% CI 22.5-33.2). The distribution of causes of death is being finalised through verbal autopsy.

#### Neural Tube Defects

Among 5,523 surveyed newborns, 0.7% were identified as having a confirmed or suspected NTD. The breakdown by type included encephalocele (20) and back swellings consistent with spina bifida or myelomeningocele (18). Microcephaly, which is not itself a neural tube defect, was observed separately in 28 newborns (0.5%) and is not included in the NTD totals. In total, 37 infants had at least one suspected or confirmed NTD (Table 3). Confirmation was pursued through follow-up visits by the CONRIM field team, with infants requiring surgical care referred to CURE Children’s Hospital or MNRH. Of the infants with confirmed NTDs in the early surveillance period, the majority had already presented to CURE Hospital independently, suggesting active care-seeking for visible defects in this population.

**Table 3.** Neural tube defects (NTDs) identified by type among enrolled newborns (N = 5,523). Suspected cases are based on Village Health Team visual screening and caregiver report; rates are per 1,000 newborns. “Any NTD” counts each infant once (≥1 positive screen). Confirmed diagnoses and CURE Children’s Hospital referrals are pending field confirmation (NA).

| NTD type | Suspected (n) | Per 1,000 newborns |
| --- | --- | --- |
| Encephalocele | 20 | 3.6 |
| Myelomeningocele (spina bifida) | 18 | 3.3 |
| **Total (sum of types)** | **38** | **6.9** |
| **Any NTD (≥1 screen positive)** | **37** | **6.7** |

Importantly, infants with anencephaly are likely underrepresented in these figures, as affected pregnancies frequently result in miscarriage or stillbirth and may not be classified as NTDs during community-level reporting. Estimates from similar settings suggest that approximately half of all infants with NTDs may present as stillbirths, indicating that the true NTD prevalence may be substantially higher than the rate captured through live-birth surveillance alone.

#### Maternal Nutrition and Folic Acid

Among mothers of enrolled newborns, 89.0% reported using any folic acid supplementation. However, the vast majority of folic acid use began after the pregnancy was confirmed rather than before conception. Pre-conception folic acid use was extremely rare, consistent with the absence of national guidelines or programmes promoting pre-conception supplementation in Uganda. The folic acid preparations available through government health centres are predominantly high-dose formulations (5 mg), prescribed primarily for anaemia management during antenatal care rather than for NTD prevention. Lower-dose formulations (500 μg), appropriate for pre-conception NTD prevention, were found to be available only at selected private pharmacies and at higher cost. These findings highlight the gap between the well-established evidence for pre-conception folic acid supplementation in NTD prevention and the current reality of supplementation practices in this population (Table 4).

**Table 4.** Maternal folic acid supplementation by timing among mothers of enrolled newborns (N = 5,523). Percentages for timing sub-categories are of mothers reporting any folic acid use.

| Folic acid variable | n | % |
| --- | --- | --- |
| **Any folic acid use** | **4,918** | **89.0%** |
| Pre-conception (before only) | 73 | 1.3% |
| During pregnancy only | 4,811 | 87.1% |
| Both before and during | 34 | 0.6% |
| No folic acid use | 605 | 11.0% |

#### Water, Sanitation and Hygiene and Environmental Risk Factors

Household WASH conditions were assessed for all enrolled households (Supplementary Table S1). The primary water source during the rainy season was borehole (45.4%) and tap water (44.9%), with smaller proportions using protected springs, shallow wells, or river or lake water. Water source distribution shifted modestly during the dry season, with a slight increase in borehole reliance (Supplementary Table S3). Latrine access was reported by 98.1% of households. Handwashing with soap was reported by 94.6% of caregivers, with the most common occasions being before handling the baby and before eating.

Cord care practices were also documented. The most common practices were applying water (40.3%) or keeping the cord dry with nothing applied (37.7%); other reported substances included baby powder (13.3%), a mixture of warm water and salt (7.7%), and, less commonly, chlorhexidine and Umbi-Gel; 0.4%. These practices are of particular interest given the established association between nonsterile cord care and neonatal sepsis risk (Supplementary Table S2).

#### Geospatial Patterns

Spatial analysis of pSBI risk across Jinja District villages, using a Bayesian hierarchical spatial model (a Besag-York-Mollié [BYM-2] model estimating both spatial and non-spatial residual variation), revealed a weak spatial trend in neonatal infection risk (Figure 4). Risk appeared modestly lower in more urban, lower-poverty areas in and around Jinja City, and no discrete hotspots were identified. These analyses excluded infants over 28 days of age and records with negative age values. Exploratory stratification by timing of infection onset suggested possible differences in geographic patterns for early-onset (<72 hours) and late-onset (≥72 hours) infections, though these analyses remain preliminary.

**Figure 4.**
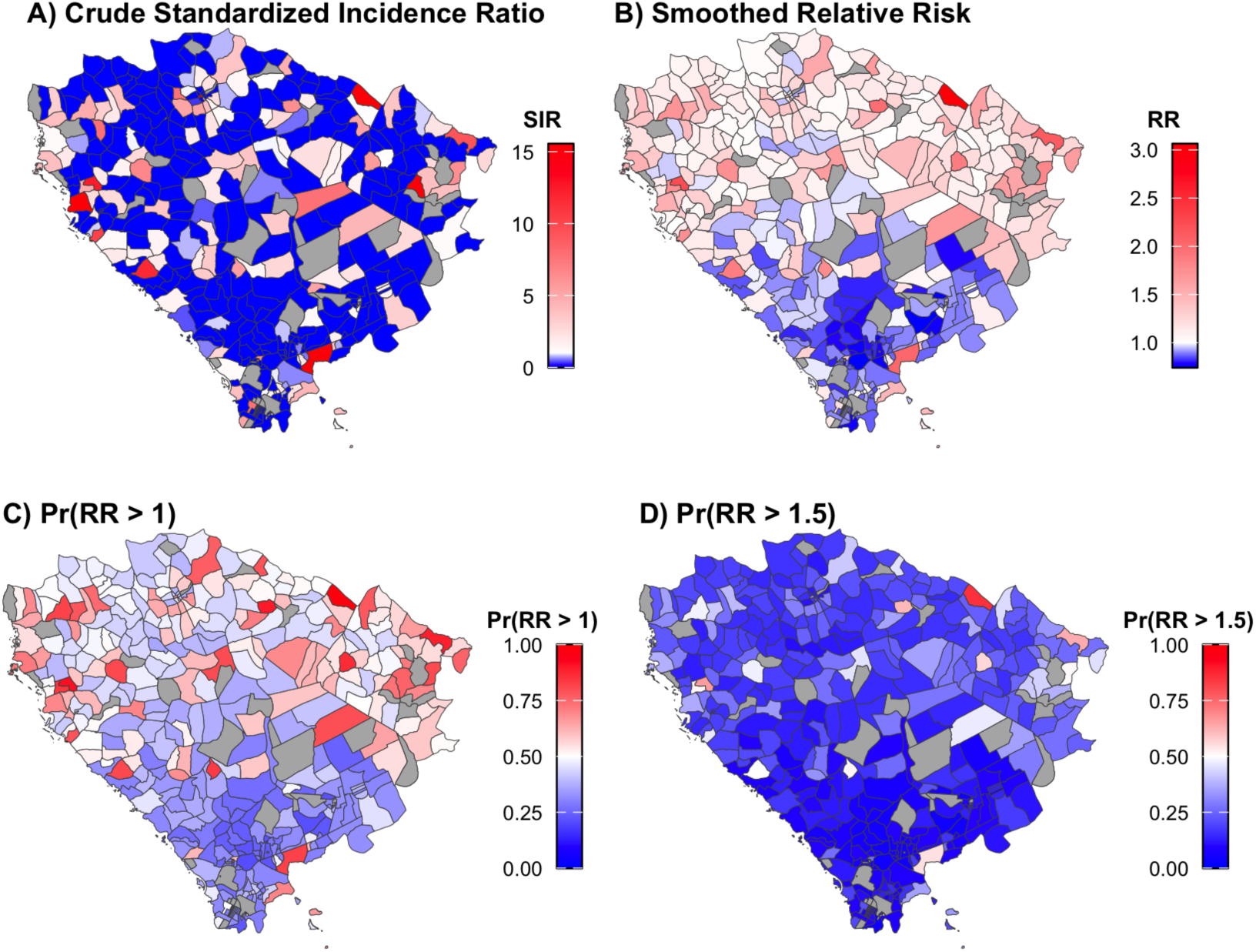
Geospatial distribution of possible serious bacterial infection (pSBI) across Jinja District, 08/21/2025-06/30/2026. A) Crude standardized incidence ratio (SIR; observed/expected pSBI counts per village), B) smoothed relative risk (RR; posterior median), and C-D) the posterior probability that a village’s RR exceeds 1 and 1.5, respectively. RR was modeled with a Besag-York-Mollié (BYM-2) spatial model using penalized-complexity (PC) priors: a weakly conservative prior on the spatial mixing parameter φ (P(φ < 0.5) = 2/3) and a moderately regularizing prior on the marginal standard deviation σ (P(σ > 1) = 0.05). Expected counts at the village level were calculated by multiplying the number of births in each village by the overall pSBI rate in Jinja District; we did not stratify on age or sex. Panel A is the crude estimate; panels B-D derive from the smoothed posterior, where B is the point estimate and C-D quantify its uncertainty. Substantial uncertainty remains. Grey polygons denote villages with no recorded births (undefined expected count). Analyses excluded infants aged >28 days at the time of survey, entries rejected on review, and entries for which families did not provide informed consent.

These preliminary findings may help inform the targeting of community health interventions and VHT resource allocation as the spatial analysis matures.

#### Microbiology Findings

Enrollment of patients with pSBI or hydrocephalus at JRRH and MNRH is in early phases. Results of blood and cerebrospinal fluid cultures and molecular diagnostics studies, along with results of environmental testing results are expected over the next few months.

## Strengths and Limitations

CONRIM’s principal strength is its population-based, community-level design, which captures births that occur outside health facilities, including home births and TBA-attended deliveries, that are systematically missed by facility-based surveillance. The universal coverage target across an entire kingdom encompassing 11 districts provides a large, geographically diverse denominator for incidence estimation. The integration of clinical, environmental, nutritional, and demographic data streams within a single platform enables multifactorial analyses of neonatal outcomes that are not possible with single-domain data sources. The real-time digital data capture system with built-in automated quality control achieves high data quality (99% clean records) while enabling rapid identification and correction of errors. The strong partnership with existing VHT infrastructure and Busoga Kingdom governance structures supports community engagement and long-term sustainability.

The multi-disciplinary PI team, spanning infectious disease, neurosurgery, neonatology, pediatrics, genomics, environmental health, nutrition, biostatistics, and public health, provides the breadth of expertise needed to pursue the platform’s integrated public health and research agendas. The longitudinal follow-up design enables ascertainment of 28-day outcomes, referral completion, and neonatal mortality, while the modular data collection system (8 forms) allows progressive enrichment of the dataset as laboratory, imaging, and surgical data become available.

This study aims to fill an important gap in the understanding of the pathogen causing neonatal sepsis. This is of vital importance because WHO recommendations for empiric antibiotic management of neonates with signs of pSBI are generalized across the whole globe and may be of limited relevance in individual locales (Cook et al., 2021; Thomson et al., 2021). National and subnational epidemiology can be used to improve the matching of empiric antibiotic therapy to the predominant local pathogens and antibiotic susceptibility patterns (Nartey et al., 2026). Additionally, targeted prevention programs aimed at reducing the most common risk factors for diseases will be more effective when based on local microbiology and infant care practices.

Similarly, efforts to reduce neural tube defects and other congenital anomalies will be more effective when the local socioeconomic and dietary practices are taken into account. Efforts to reduce NTDs through food fortification are likely to fail in communities where the majority of food is grown at home rather than purchased. Additionally, the safety of wide-scale folate supplementation in malaria-endemic areas has never been demonstrated and a careful weighing of the risks and benefits needs to be undertaken. CONRIM aims to address these considerations and quantify the risks of NTDs, maternal malaria infection and its complications and the acceptability of folate supplementation in women of childbearing age.

Several limitations should be acknowledged. VHT-reported danger signs are based on training in recognition of clinical signs but are not confirmed by a clinician at the point of capture; the sensitivity and specificity of VHT-based pSBI screening relative to clinical diagnosis have not yet been formally evaluated in this cohort. Similarly, NTD screening relies on visual inspection and caregiver report, and not all infants with suspected NTDs have been confirmed by specialist examination at the time of this report. Gestational age assessment is based on maternal recall without ultrasound dating in most pregnancies, introducing imprecision. Antibiotic use before clinical evaluation, which is common in this setting, may obscure the true incidence of culture-confirmed sepsis. Laboratory confirmation of neonatal sepsis is facility-based and not yet fully operational across all referral sites. Follow-up completion rates are still improving as the telephone follow-up system reaches full capacity. Some subcounties have lower VHT engagement and reporting completeness than others, and non-consent, while low and declining, introduces potential selection bias. Cultural barriers, including reluctance to discuss infant deaths and reliance on traditional healing practices, may lead to underreporting of neonatal morbidity and mortality.

## Collaboration and Data Access

CONRIM welcomes collaboration with researchers and public health practitioners working on neonatal health, infectious disease, birth defects, maternal nutrition, environmental health, and related fields. External investigators interested in proposing analyses using CONRIM data should contact the corresponding author to discuss the proposed research question, required data elements, and authorship arrangements. All proposals are reviewed by the CONRIM PI Group for scientific merit, feasibility, overlap with ongoing work, and compliance with ethical and institutional requirements. Data sharing is subject to the consortium’s data governance policies and relevant IRB and institutional agreements. The CONRIM PI Charter and authorship framework govern the process for all shared outputs.

## Future Directions

The CONRIM platform is designed to support a growing portfolio of research and public health activities. Near-term priorities include the scale-up of laboratory-based sepsis confirmation across all referral sites, deployment of portable MRI neuroimaging for NTD and hydrocephalus characterisation, and expansion of the 28-day follow-up system to improve outcome ascertainment. The identification of geospatial hotspots of neonatal infection risk will inform the design of targeted community-level interventions, including folic acid supplementation campaigns, cord care education, and referral pathway strengthening.

Medium-term objectives include genomic studies of folate metabolism pathway variants and NTD susceptibility, environmental risk factor analyses linking household conditions and satellite-derived environmental exposures to neonatal outcomes, integration with national Health Management Information System data for external validation of surveillance completeness, and verbal autopsy standardisation for neonatal deaths. The platform’s infrastructure, including the ODK data pipeline, real-time dashboards, and VHT network, is designed to be extensible and could support future interventional studies, vaccine trials, or implementation research within the established cohort.

Ultimately, CONRIM aims to generate the population-level evidence needed to inform national and regional policy on neonatal health, folic acid fortification, infection prevention, and health system strengthening in Uganda and across sub-Saharan Africa. The platform’s emphasis on capacity building and local research leadership is intended to ensure that the surveillance infrastructure and the expertise it generates are sustainable beyond the initial funding period.

## Patient and Public Involvement

Community engagement is central to CONRIM’s design and operations. The VHT members who serve as the primary data collection workforce are themselves community residents who volunteer to serve in the VHT role, providing a direct link between the study and the populations it serves. VHT leaders participate in regular community meetings where study progress, preliminary findings, and operational challenges are discussed. The Busoga Kingdom’s traditional governance structures have been engaged as partners, and their endorsement has facilitated community acceptance and trust. Mothers and caregivers enrolled in the study receive feedback through the follow-up system, including health education on neonatal danger signs, referral guidance for sick infants, and information on cord care and breastfeeding practices. Study participation cards issued to enrolled households serve both as a data linkage tool and as a tangible acknowledgement of participation. The study team has adapted questionnaire language, consent processes, and follow-up scripts in response to community feedback, including simplification of technical terms and addition of visual aids for VHTs and respondents with limited literacy.

## Funding

CONRIM is supported by the Ruddy Lifesaving Fund at Yale University. Additional support for development of rapid antigen testing for *P. thiaminolyticus* is from NIH grant R21AI196636 (JRB).

## Ethics Statements

### Ethics approval

This study was approved by the Yale University Institutional Review Board (protocol #2000040126, approved 30 July 2025) and by Mbarara University of Science and Technology Research Ethics Committee (MUST-REC, protocol #20241657, approved 17 January 2025). Informed consent was obtained from all participating caregivers.

## Competing Interests

None declared.

## Author Contributions

JNP, JEE, and SJS conceived and designed the study and provided supervision. JNP, AJW, ST, and JH developed the methodology, analyzed and interpreted the data, and drafted the manuscript. JH, RM, KR, and MOs contributed to data management, analysis, software, validation, and visualization. DN, KS, MOc, EMBK, BKN, POO, AK, NJ, HM, HN, AN, S, IT, AY, MJ, EKi, AKa, JA, HA, JBT, and EO contributed to investigation, data collection, resources, and project administration. JRB, SUM, JM, AM, and EK contributed to scientific oversight, resources, and critical revision of the manuscript. SJS contributed to funding acquisition. All authors reviewed and approved the final manuscript.

## Data Availability

Data from the CONRIM surveillance platform are available to external researchers through a structured proposal process. Enquiries should be directed to the corresponding author. Data sharing is subject to the consortium’s data governance policies and relevant IRB and institutional agreements.

## Acknowledgements

VHTs and VHT leadership across the district; Busoga Kingdom traditional governance; JRRH and Mulago staff; CURE Children’s Hospital of Uganda; Makerere University; field supervisors and data management team; study participants and their families. And special thanks to the King (Kyabazinga), Queen (Inhebantu) and Prime Minister (Katikuru) of Busoga.

## Supplementary Material

## Supplementary Tables

**Supplementary Table S1.** Household water, sanitation and hygiene (WASH) indicators (N = 5,523). Values are n (%). Improved water source reflects borehole or tap water during the rainy season.

| Indicator | n (%) |
| --- | --- |
| Improved water source (borehole/tap, rainy) | 4,988 (90.3%) |
| Latrine access | 5,418 (98.1%) |
| Handwashing with soap | 5,226 (94.6%) |

**Supplementary Table S2.** Substances applied to the umbilical cord stump (N = 5,523). Values are n (%); categories are not mutually exclusive where more than one substance was reported.

| Substance | n | % |
| --- | --- | --- |
| Water | 2,228 | 40.3% |
| Nothing applied (kept dry) | 2,080 | 37.7% |
| Baby powder | 737 | 13.3% |
| Water and salt | 424 | 7.7% |
| Alcohol / spirits | 86 | 1.6% |
| Other (specified) | 66 | 1.2% |
| Petroleum jelly | 51 | 0.9% |
| Other | 38 | 0.7% |
| Chlorhexidine (Umbi-gel) | 21 | 0.4% |
| Herbs | 23 | 0.4% |

**Supplementary Table S3.** Primary household water source by season (N = 5,523). Values are n (%) within each season.

| Water source | Rainy season n (%) | Dry season n (%) |
| --- | --- | --- |
| Borehole | 2,507 (45.4%) | 2,784 (50.4%) |
| Tap water | 2,481 (44.9%) | 2,135 (38.7%) |
| Protected source | 331 (6.0%) | 407 (7.4%) |
| Shallow well | 111 (2.0%) | 111 (2.0%) |
| Unprotected well | 43 (0.8%) | 48 (0.9%) |
| Harvested rainwater | 28 (0.5%) | 13 (0.2%) |
| Other | 22 (0.4%) | 25 (0.5%) |

## Supplementary Figures

**Supplementary Figure S1.**
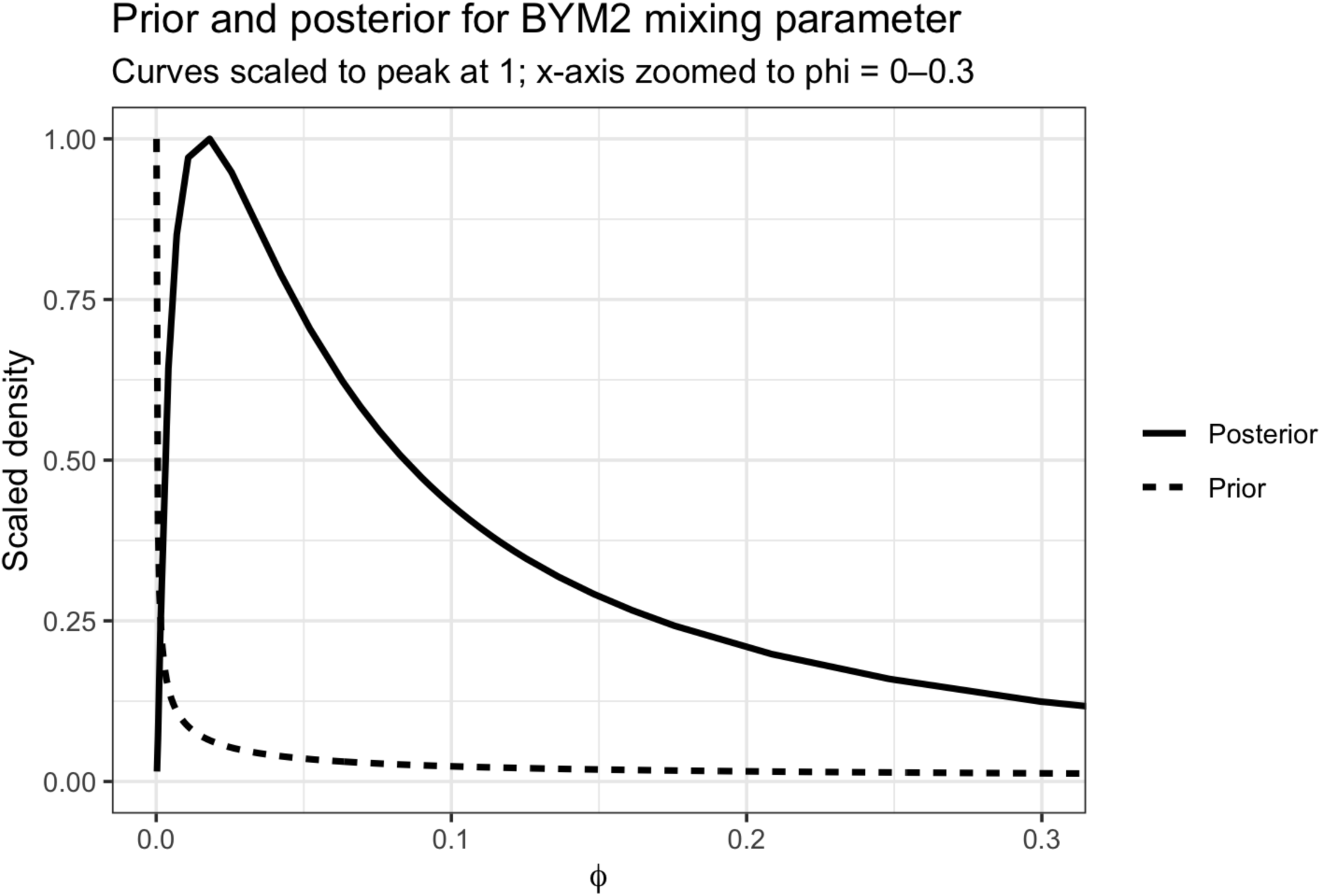
Prior and posterior for the BYM-2 spatial mixing parameter. Prior (dashed) and posterior (solid) for φ, the proportion of the random-effect variance that is spatially structured. The penalized-complexity prior was weakly conservative toward the unstructured model (P(φ < 0.5) = 2/3). Curves are scaled to a common peak of 1 and the x-axis is truncated at φ = 0.3 for visibility; heights therefore reflect shape, not probability mass.

**Supplementary Figure S2.**
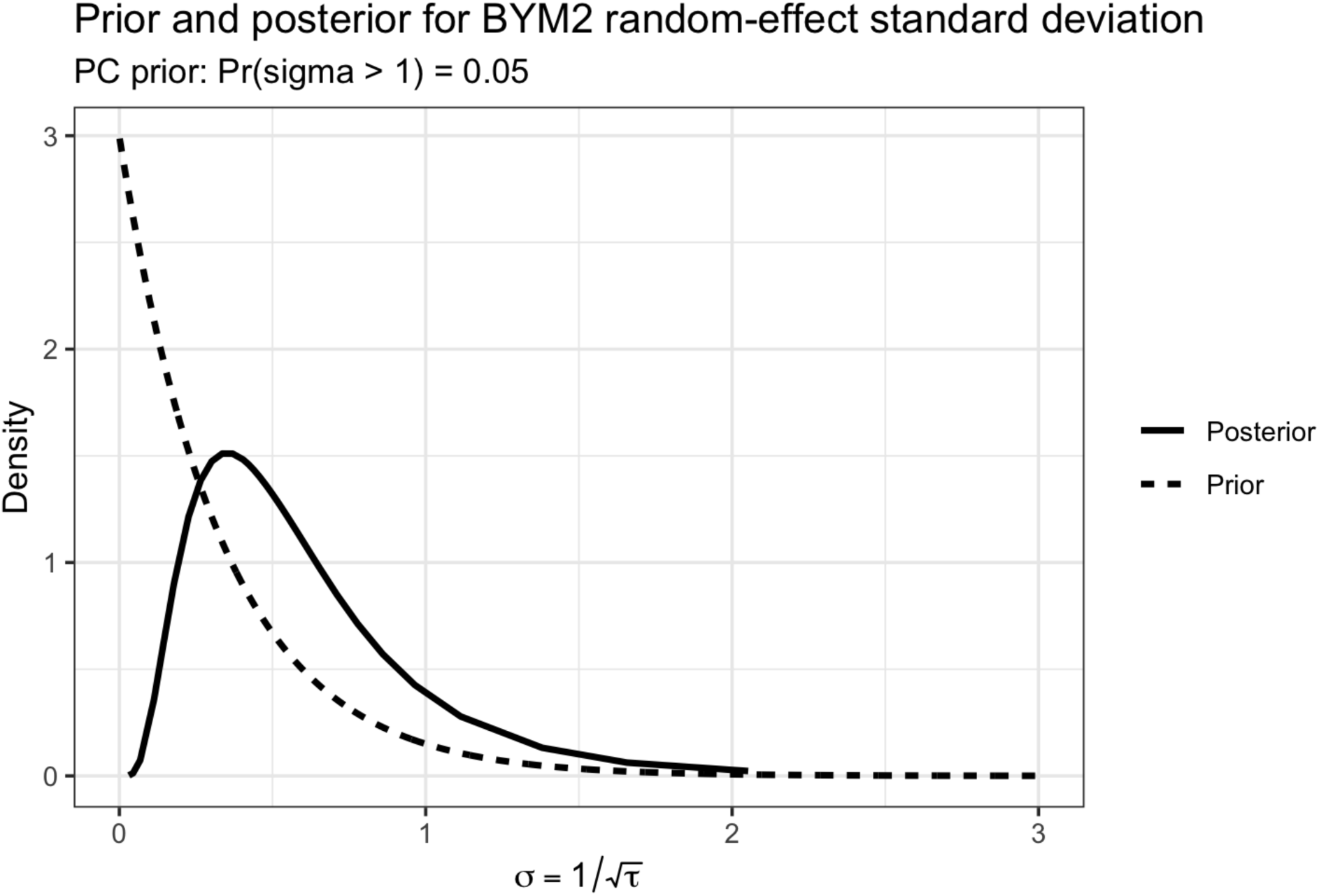
Prior and posterior for the BYM-2 random-effect standard deviation. Prior (dashed) and posterior (solid) for σ = 1/√τ, the marginal standard deviation of the combined BYM-2 random effect on the log-RR scale. The penalized-complexity prior is Exponential with P(σ > 1) = 0.05. The posterior concentrated around a mode of ∼0.4, well inside the prior and clearly data-informed, indicating a modest overall magnitude of village-level variation in pSBI risk.

## Notes

### Competing Interest Statement

The authors have declared no competing interest.

### Author Declarations

The study was approved by the Yale University Institutional Review Board (protocol #2000040126, approved 30 July 2025) and has received local ethical review and administrative clearance from the Research Ethics Committee of the Mbarara University of Science and Technology, the Uganda National Council for Science and Technology, and relevant district authorities (protocol #MUST-2024-1657, approved 8 August 2024).

